# Transcriptional aging across CNS tissues in the ALS-FTD spectrum reflects clock architecture and disease-associated cellular signatures

**DOI:** 10.64898/2026.11.19.26368520

**Authors:** Tianying Zhao, Gang Wu, Mariah M. Hoffman, Veronique V. Belzil

## Abstract

**Background:** We examined transcriptomic (gene expression-based) aging in postmortem central nervous system (CNS) tissues from donors across the amyotrophic lateral sclerosis (ALS)-frontotemporal dementia (FTD) spectrum and investigated the molecular and cellular features underlying disease-associated differences in transcriptional aging clocks.

**Methods:** Six transcriptomic clocks (RNAAgeCalc DESeq2, GTExAge, Pearson and GenAge; Pasta REG and age-shift) were applied to 2,235 RNA-sequencing samples from 609 donors across 11 CNS regions, with adjustment for age and tissue-specific technical variables. We compared transcriptional age across ALS, ALS-FTD, FTD, and controls, examining regional variation, gene-level contributions, cell composition, disease duration, and *C9orf72* status, with independent validation of gene contributions in a cervical spinal cord cohort.

**Results:** Disease-associated differences in transcriptional age varied by diagnosis, tissue, and clock. ALS and ALS-FTD showed the strongest differences in spinal cord, while FTD differences localized primarily to the frontal and temporal cortex. In ALS spinal cord, RNAAgeCalc DESeq2 estimates were lower than controls, whereas other clocks estimated higher transcriptional age when significant. *CHIT1* was identified as the primary contributor to the DESeq2 transcriptional age decrease in ALS. Cell composition adjustment attenuated ALS spinal cord differences, reflecting microglial, lymphocyte, oligodendrocyte, and astrocyte signatures, though the lumbar GenAge increase in transcriptional age and DESeq2 decrease remained significant. The higher transcriptional age in ALS motor cortex also remained significantafter adjustment. Clinical associations were likewise clock dependent. Higher spinal DESeq2 age acceleration (AA) was associated with longer ALS duration, though this association was strongly influenced by the *CHIT1* term. Higher GTExAge AA in cervical spinal cord, thoracic spinal cord, and occipital cortex was associated with shorter disease duration; no significant associations were observed for the other clock–tissue combinations or for ALS-FTD. No clock differed by *C9orf72* repeat expansion status.

**Conclusions:** In this study we found disease-associated differences in transcriptional age that localize to clinically relevant tissues. Differences in transcriptional aging across the ALS-FTD spectrum reflect interactions among clock architecture, disease-associated gene expression, tissue region, and cellular composition. Resolving these components reveals the biological context underlying divergent clock estimates and provides a framework for interpreting changes in transcriptional aging.

## 1. Introduction

Amyotrophic lateral sclerosis (ALS) is characterised by progressive loss of voluntary motor function (Vasta et al. 2025), whereas frontotemporal dementia (FTD) encompasses a group of neurodegenerative diseases characterised by progressive deficits in behaviour, executive function or language (Bang et al. 2015). Despite their distinct clinical presentations, ALS and FTD form an overlapping disease spectrum. Individuals with ALS may develop cognitive and behavioural impairment, including frontotemporal dementia (ALS-FTD), while individuals presenting with FTD may also develop motor neuron dysfunction (Strong et al. 2017). This overlap extends to their genetic architecture, with *C9orf72* repeat expansions representing a shared genetic cause of ALS and FTD identified independently in families with FTD, ALS, and combined FTD/ALS phenotypes (DeJesus-Hernandez et al. 2011; Renton et al. 2011). Studying ALS, ALS-FTD and FTD together therefore provides an opportunity to examine shared and disease-specific molecular patterns.

Across the ALS-FTD disease spectrum, age is a major risk factor. Consistent with this observation, population-based studies from European ALS registries have demonstrated an approximately linear relationship between log incidence and log age (Al-Chalabi et al. 2014). FTD has historically been considered an early-onset dementia, but its incidence also increases with age, with a substantial disease burden occurring after age 65 (Urso et al. 2025). Nevertheless, substantial heterogeneity in age at onset remains, and currently identified genetic, environmental, and lifestyle risk factors explain only part of this variation (Vasta et al. 2022). Several pathogenic mechanisms implicated in ALS/FTD overlap with pathways that are also involved in the aging process, including autophagy, inflammation, nucleocytoplasmic transport, and RNA splicing (Niccoli et al. 2017). These observations motivate investigation of molecular measures of aging that may capture biological variation beyond chronological age alone.

Chronological age is the standard reference for aging studies, but it does not fully describe interindividual variation in biological aging (Min et al. 2024). Biological age is an abstract construct that reflects an individual’s overall health state and underlying aging processes that cannot be directly observed or quantified (Kriukov et al. 2025). Alterations in gene expression have been associated with aging and age-related outcomes (de Magalhães et al. 2009; Tacutu et al. 2018; Welle et al. 2003; Rodwell et al. 2004; Lu et al. 2004; Glass et al. 2013; Peters et al. 2015; Fleischer et al. 2018). RNAAgeCalc is a multi-tissue transcriptional age calculator developed using RNA-sequencing data from the Genotype-Tissue Expression (GTEx) project and provides both across-tissue and tissue-specific age predictors (Ren and Kuan 2020). Pasta (Predicting Age-Shift from Transcriptomic Analysis) is a more recent multi-tissue transcriptomic aging clock, trained on 17,212 healthy samples from 21 studies using an "age-shift" learning framework, that predicts relative age across tissues and data types, including bulk and single-cell RNA-sequencing and microarray data (Salignon et al. 2026). Transcriptional age acceleration (AA) quantifies the deviation of estimated transcriptional age from that expected for chronological age and may therefore capture biologically meaningful variation in transcriptional aging.

Transcriptomic profiling has increasingly been used to characterize molecular heterogeneity in ALS and individual differences in brain aging. Using postmortem motor cortex gene expression data, Marriott et al. identified three molecular phenotypes linked to ALS pathogenesis: synaptic and neuropeptide signaling, oxidative stress and apoptosis, and neuroinflammation. RNAAgeCalc-derived transcriptional AA was significantly lower in cluster 2 than cluster 1 in TargetALS motor cortex, with a similar but nonsignificant trend in KCL BrainBank, whereas no significant differences across clusters were observed in either blood dataset (Marriott et al. 2023). Using transcriptome-wide cerebral cortex gene expression data from several cohorts totaling 1,904 autopsied human brain samples, Rhinn and Abeliovich quantified individual variability in age-associated phenotypes and identified *TMEM106B* and *GRN* loci as determinants of differential aging in the cerebral cortex (Rhinn and Abeliovich 2017).

Transcriptomic clocks have not yet been compared systematically across the ALS-FTD disease spectrum or across multiple central nervous system (CNS) tissues, including the spinal cord, where lower motor neurons degenerate in ALS. Clock estimates from bulk tissue may also reflect disease-related changes in gene expression and cell composition, as well as aging. In this study, we applied six transcriptomic clocks to 2,235 postmortem RNA-sequencing samples from 609 donors across 11 CNS regions in the New York Genome Center (NYGC) ALS Consortium cohort (Humphrey et al. 2026), including the RNAAgeCalc DESeq2, GTExAge, Pearson and GenAge signatures (Ren and Kuan 2020) and the Pasta regression and age-shift models (Salignon et al. 2026). We compared adjusted transcriptional age in ALS, ALS-FTD and FTD relative to non-neurological controls in each tissue and compared transcriptional age across tissues within each disease group. To identify the contributors to those differences, we decomposed clock estimates into individual gene contributions, evaluated these contributions in an independent cervical spinal cord dataset, and adjusted for cell composition estimated from the bulk RNA data. Finally, we examined associations between transcriptional age acceleration and ALS disease duration and *C9orf72* repeat expansion status.

## 2. Methods

### 2.1 Data and study population

We analysed NYGC ALS Consortium maximum likelihood expected counts and metadata (GEO GSE137810; Zenodo 18989237 and 20090702; Supplementary Table S1). Raw downloaded data included 58,929 genes across 2,574 samples preprocessed using the RAPiD-nf framework (STAR 2.7.2a, RSEM 1.3.1, GRCh38/GENCODE v30) (Di Tommaso et al. 2017; Humphrey et al. 2026). X/Y counts were summed for 45 pseudoautosomal genes. For this study, chronological age was defined as age at death. Donors whose age was reported only as “90 or older” were treated as having missing age and excluded from analyses requiring age at death; this excluded three control donors (15 samples). No ALS, ALS-FTD, or FTD donors were excluded for this reason. For the one donor with conflicting values, age at death was retained because it matched age at onset plus disease duration. *C9orf72* repeat expansion status was taken from the consortium’s annotation, based on whole-genome sequencing and repeat-primed polymerase chain reaction. Donors with ALS, ALS-FTD, FTD or no known neurological disease (controls) were analysed. For the four donor–tissue pairs sequenced twice, we retained the sample with the higher RNA integrity number (RIN), using read count to break ties. After deduplication and removal of samples with missing age or covariate data, the analysis included 2,235 samples from 609 donors across 11 regions with ≥50 samples each. Analyses were performed using Python and R.

### 2.2 Transcriptomic clocks

We used six published transcriptomic aging clocks: four brain-specific predictors implemented in RNAAgeCalc v1.22.0, based on the DESeq2 (472-gene), GTExAge (485-gene), Pearson (411-gene), and GenAge (302-gene) feature sets (Ren and Kuan 2020), and the 8,113-gene Pasta regression (REG) and age-shift models (Salignon et al. 2026). RNAAgeCalc transcriptional age equalled intercept + Σ w_g_ log (FPKM_g_ + 1), where w_g_ is the gene weight and FPKM is fragments per kilobase per million mapped reads. Only the RNAAgeCalc DESeq2 model contained *CHIT1* as a feature. Clock units were years of transcriptional age (RNAAgeCalc), years of predicted age (Pasta REG) and age-score years (Pasta age-shift; one unit per year of age difference).

For each clock and tissue, Pearson correlation between transcriptional age and age at death was calculated separately in two groups: control donors and patients (ALS, ALS-FTD and FTD pooled). P values were corrected across the 11 tissues using Benjamini–Hochberg false discovery rate (FDR), separately for each clock and group (Benjamini and Hochberg 1995). Unadjusted linear fits were used for visualization.

### 2.3 Association of disease status with transcriptional age acceleration

Each disease group was compared with controls in each tissue and for each clock by linear regression:

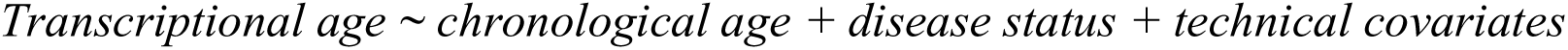

The disease-status coefficient therefore represented the difference in transcriptional age relative to controls after adjustment for chronological age. Technical covariates were the tissue-specific covariates used in the consortium’s differential expression models, selected using an empirical strategy (Methods Table 3; (Humphrey et al. 2026)). The consortium provided no covariates for occipital cortex, which was therefore adjusted for library preparation, the covariate shared by nine of the ten consortium tissue models. Tissues with fewer than eight samples in either group were not tested. FDR was computed across the tested tissues within each disease group and clock.

### 2.4 Tissue contrasts in transcriptional age within disease groups

Transcriptional age of each clock was fitted separately within ALS, ALS-FTD and FTD using linear mixed models with donor as a random intercept:

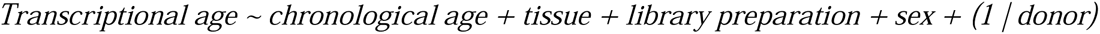

Each model included samples with complete chronological age, sex and library preparation, from tissues with at least eight such samples in the disease group and no more than one sample per donor-tissue pair. Models were fit with restricted maximum likelihood with cerebellum as the reference tissue. Tissue contrasts were evaluated using Wald tests (z = estimate/SE) and normal approximation 95% confidence intervals (CIs) (estimate ± 1.96 × SE), where SE denotes the standard error. P values were FDR-adjusted across tissue contrasts within each disease group and clock. Because transcriptional age was the outcome, positive tissue coefficients indicate higher adjusted transcriptional age relative to cerebellum, in the clock-specific units defined above (Methods 2.2).

### 2.5 Gene contributions and independent validation

To quantify the contribution of individual genes to the ALS–control difference in RNAAgeCalc DESeq2 transcriptional age, we multiplied each gene’s fixed clock weight by its adjusted ALS– control difference in log_2_(FPKM + 1), estimated using the model described in Methods 2.3. Gene-level contributions were expressed in years of transcriptional age and summed to the overall ALS–control clock difference. We separately estimated ALS–control differences for the full 472-gene DESeq2 clock, the *CHIT1* term, and the remaining 471-gene component using the same model. FDR correction was applied across tissues separately for each outcome.

We assessed reproducibility of these gene-level contributions in an independent cervical spinal cord RNA-seq cohort comprising 7 donors with sporadic ALS and 8 controls (SRA SRP064478, processed through recount3) (Brohawn et al. 2016; Wilks et al. 2021). Given the smaller validation sample numbers, ALS–control expression differences were estimated using parsimonious models adjusted for chronological age only, and the resulting gene contributions were compared with those from the primary cohort.

Finally, to assess whether increased *CHIT1* expression in ALS spinal cord could be explained by altered microglial abundance, we modeled log_2_(FPKM + 1) *CHIT1* expression in cervical and lumbar spinal cord as a function of ALS status, chronological age and technical covariates, first without and then with estimated microglial fraction. Microglial effects were expressed per 10-percentage-point increase in estimated fraction, with FDR correction across the two spinal cord levels separately for each model term.

### 2.6 Cell composition

Cell type fractions estimated using Multi-subject Single Cell deconvolution (MuSiC) (Wang et al. 2019) were available from the ALS consortium for 11 spinal cord, eight cortical, and 13 cerebellar cell types in ALS and non-neurological control tissues. The consortium supplied no estimates for occipital cortex, hippocampus, thoracic cord or ALS-FTD/FTD samples, so these were not composition-adjusted.

Following the compositional approach of Merbaum *et al*., modeling included all estimated cell type fractions except one reference cell type to avoid collinearity. Within each tissue, the reference was the cell type with the highest mean and standard deviation across eligible samples. (Merbaum et al. 2025). The reference cell type was oligodendrocytes in all tissues except temporal cortex, where excitatory neurons were used, and cerebellum, where granule cells were used. The reference cell type was chosen once per tissue and used in every model.

Linear models with the Methods 2.3 covariates compared cell type fractions between ALS and controls:

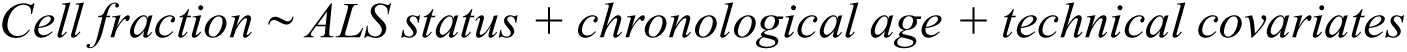

Clock models in spinal cord and unspecified motor cortex were compared before and after composition adjustment using identical samples, with model fit assessed by adjusted R², Akaike Information Criterion, and partial F-tests. Fractions were expressed as percentages, so differences are in percentage points, with FDR across all tested tissue-cell-type comparisons. To identify the cell types contributing to each ALS–control difference, the Methods 2.3 model was fitted on the composition-available samples without cell fractions and again with each cell type fraction individually, as well as with all fractions except the reference:

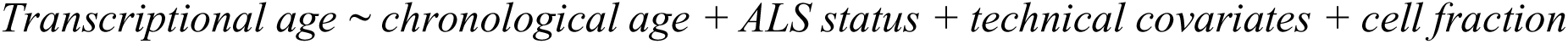

ALS−control differences with and without each fraction were compared, with FDR across tissues within clock and adjustment. This analysis was restricted to samples with cell composition estimates and used FDR correction across the eight eligible tissues; therefore, statistical significance may differ from that in Methods 2.3.

### 2.7 Disease duration

Transcriptional AA was calculated within each tissue as the residual of transcriptional age regressed on chronological age and the tissue’s technical covariates (Methods 2.3), using all analysis-set samples within that tissue including disease groups and controls:

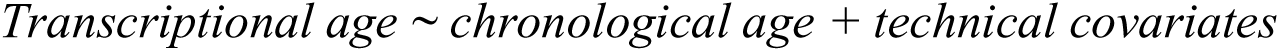

AA was calculated for each clock and for RNAAgeCalc DESeq2 transcriptional age excluding the *CHIT1* contribution (other 471 genes). *CHIT1* expression, in log (FPKM + 1), was adjusted with the same model:

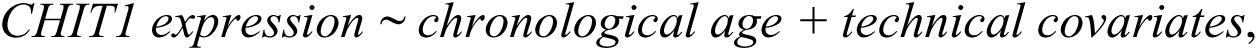

and the model residuals were the adjusted *CHIT1* expression.

Onset-to-death duration was analysed separately within ALS and ALS-FTD, for each tissue with ≥ 30 donors with complete data (11 ALS tissues and four ALS-FTD tissues). Exposures were AA and adjusted *CHIT1* expression. Each exposure entered a separate Cox proportional hazards model with the consortium’s case-only covariates (Humphrey et al. 2026):

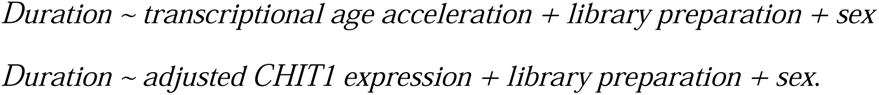

The hazard ratio (HR) was per AA year (RNAAgeCalc and Pasta REG), per age-score year (Pasta age-shift) or per log_2_ unit (*CHIT1*). An HR <1 indicates a lower instantaneous hazard of death and therefore longer disease duration, whereas an HR >1 indicates a higher hazard and shorter disease duration. Because the *CHIT1* term equals its fixed weight (−4.33) × expression, adjusted expression equals *CHIT1* term AA divided by −4.33. FDR correction was applied across tissues within each disease group, clock, and exposure.

### 2.8 *C9orf72* status

Each clock was compared between ALS *C9orf72* repeat expansion carriers and non-carriers within each tissue, adjusting for chronological age, library preparation and sex, then additionally for cell composition except the reference type. Seven tissues were analysed; tissues with fewer than eight carriers or non-carriers were excluded. FDR correction was applied across the seven tissues, separately for each clock and for models with and without cell composition.

## 3. Results

For this analysis, we utilized publicly available bulk RNA-sequencing data from the New York Genome Center ALS Consortium resource (Humphrey et al. 2026). Data were deduplicated so that each donor contributed no more than one sample per tissue. The final cohort included 2,235 samples from 609 donors across 11 central nervous system regions, each represented by at least 50 samples. Donor characteristics are summarised by disease group (Table 1).

**Table 1.**
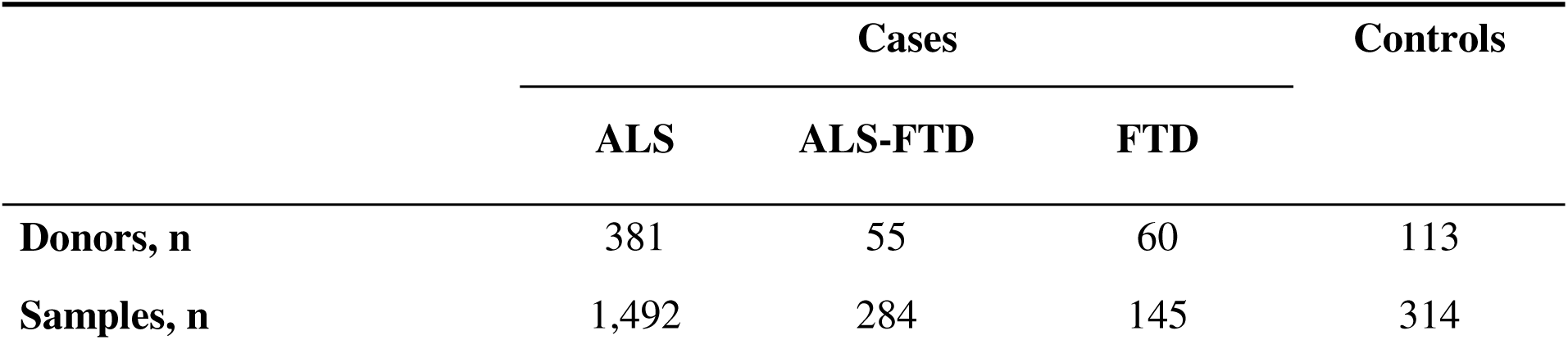
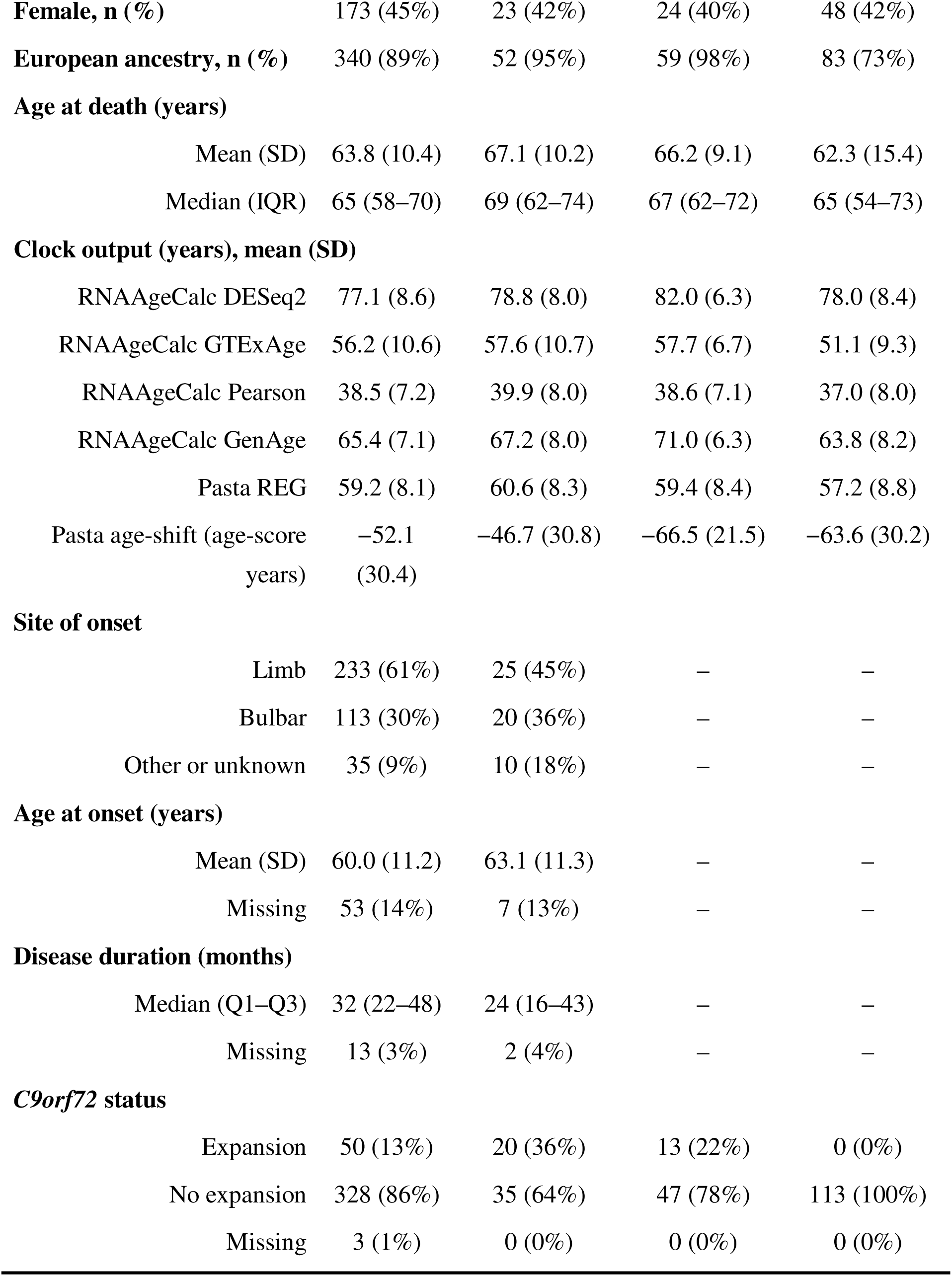
Characteristics of the study population by disease group.

Values are donor n (%) unless stated otherwise. Clock outputs are sample means (SD), one per donor/tissue, pooled across tissues and therefore dependent on tissue mix; RNAAgeCalc/Pasta REG, years; Pasta age-shift, age-score years. Duration, onset to death; all donors had died. -, not recorded; IQR, interquartile range; Q1-Q3, first-third quartile.

### 3.1 Transcriptomic clocks correlate with chronological age in most tissues

Prior to modeling differences in transcriptional age between cases and controls, we examined the correlation of transcriptional age and chronological age in the ALS consortium cohort. Correlations were calculated separately in non-neurological controls and in patients (ALS, ALS-FTD and FTD pooled). Of the 66 clock–tissue combinations, 53 in controls and 62 in patients showed significant correlations between transcriptional age and chronological age after FDR adjustment, with moderate to moderately high Pearson’s r values (Supplementary Figure S1). In the occipital cortex, no correlation for the control cohort (n = 11) reached significance after adjustment. Correlations were also low across all the RNAAgeCalc clocks (mean r = 0.26) and only moderate across the Pasta clocks (mean r = 0.49). In the thoracic spinal cord, which had the fewest control samples (n = 8), the correlation between chronological age and transcriptional age failed to reach nominal significance in 3 of 4 RNAAgeCalc clocks, but correlations remained moderately high across the RNAAgeCalc clocks (mean r = 0.69) and Pasta clocks (mean r = 0.75). No clear evidence of non-linearity was observed in either the non-neurological control or pooled patient group in the relationship between chronological age and transcriptional age. Chronological age was therefore modeled as a linear term.

Among the clocks, RNAAgeCalc Pearson and GenAge were not significantly correlated with chronological age in occipital cortex or thoracic spinal cord. The Pearson clock was additionally non-significant in lateral and medial motor cortex, and GenAge in temporal cortex and unspecified motor cortex. RNAAgeCalc DESeq2 was also found to overestimate age and show compression of the predicted age range. Correlations with chronological age were higher in controls than in patients for 54 of 66 clock–tissue combinations.

### 3.2 Disease-associated differences in transcriptional age localize to clinically relevant tissues

We modeled the association between transcriptional age and disease status, adjusting for chronological age and relevant technical covariates. With non-neurological controls as the reference group, the disease status coefficient represents the adjusted disease-control difference in transcriptional age; positive values indicate higher transcriptional age in the disease group relative to controls at the same chronological age, or, for the Pasta age-shift model, higher age-score years.

In ALS, disease status was significantly associated with differences in transcriptional age/age-score across all six clocks in cervical spinal cord, five clocks in lumbar spinal cord, four clocks in unspecified motor cortex, and one clock in thoracic spinal cord (Figure 1). No other tissues showed a significant association with disease after adjustment. All significant disease-associated coefficients indicated higher adjusted transcriptional age/age-score years relative to controls, except RNAAgeCalc DESeq2 in the cervical and lumbar spinal cord, which indicated lower transcriptional age.

**Figure 1.**
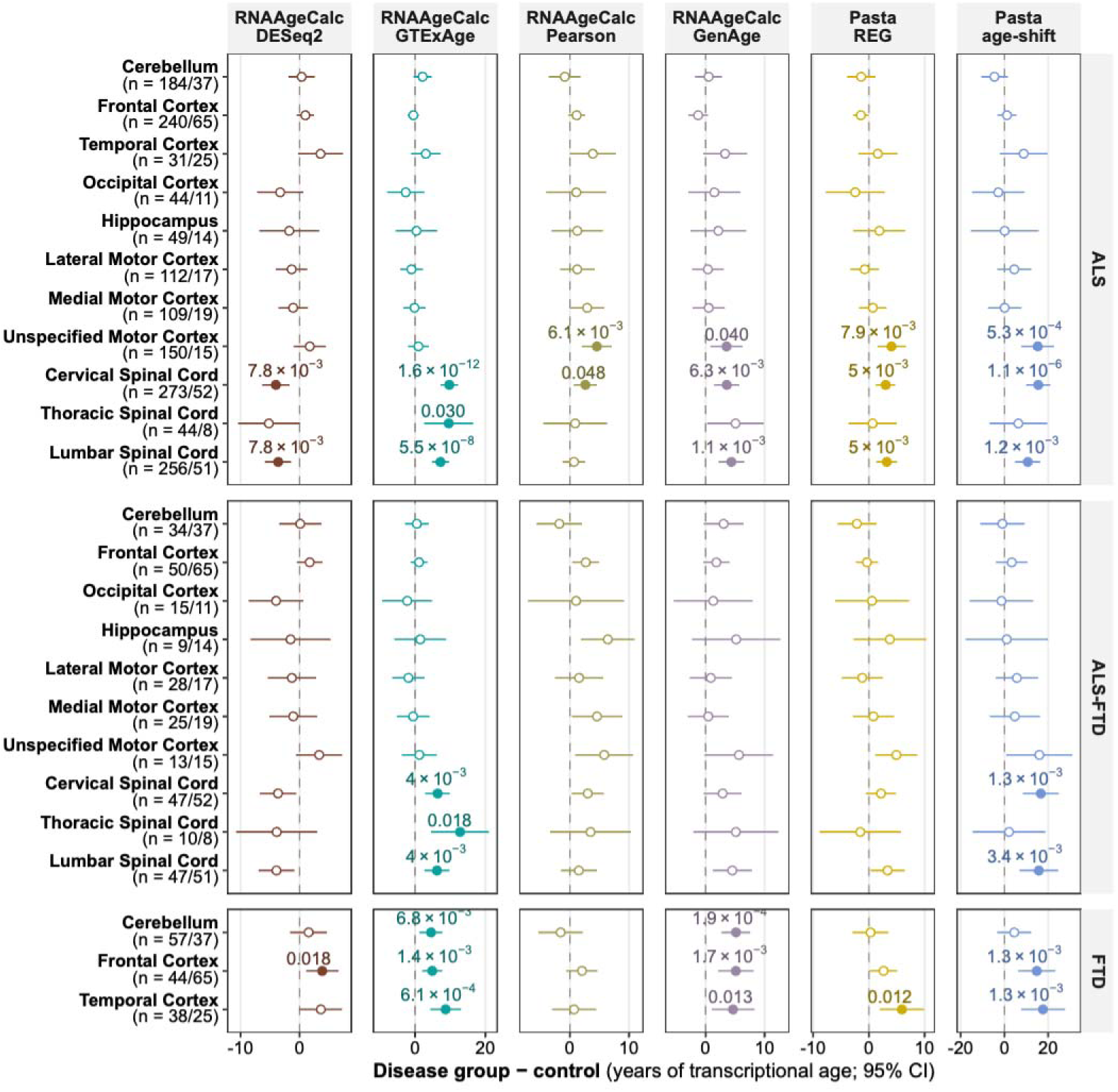
Associations between disease and transcriptional age across six transcriptomic clocks in ALS, ALS-FTD and FTD, by tissue. Disease group–control differences (95% CIs) for ALS (top), ALS-FTD (middle) and FTD (bottom) in tissues with ≥ 8 cases and controls. Linear models were adjusted for age and tissue-specific technical covariates. Units are years of transcriptional age or age-score years (Pasta age-shift). Filled points and labels indicate FDR < 0.05 across tissues within disease group and clock; n, cases/controls.

Similarly, ALS-FTD was associated with higher adjusted transcriptional age, including significant increases in Pasta age-shift scores in cervical and lumbar spinal cord and GTExAge scores in cervical, thoracic, and lumbar spinal cord. For FTD, only three tissues had sufficient sample sizes to meet inclusion criteria: cerebellum, frontal cortex, and temporal cortex. Significant increases in transcriptional age were observed in frontal and temporal cortices for four models and in cerebellum for two models. All significant disease-associated coefficients indicated higher adjusted transcriptional age/age-score years relative to controls.

### 3.3 Transcriptional age differs systematically across tissues within ALS, ALS-FTD and FTD

We compared transcriptional age across tissues within each disease group using linear mixed models that accounted for repeated tissues from the same donor (Figure 2). Cerebellum was selected as the reference tissue because it has shown comparatively limited disease-associated transcriptional alterations relative to cortical and spinal cord regions in the NYGC ALS cohort (Humphrey et al. 2026). Our disease-control analyses were broadly consistent with this regional pattern.

**Figure 2.**
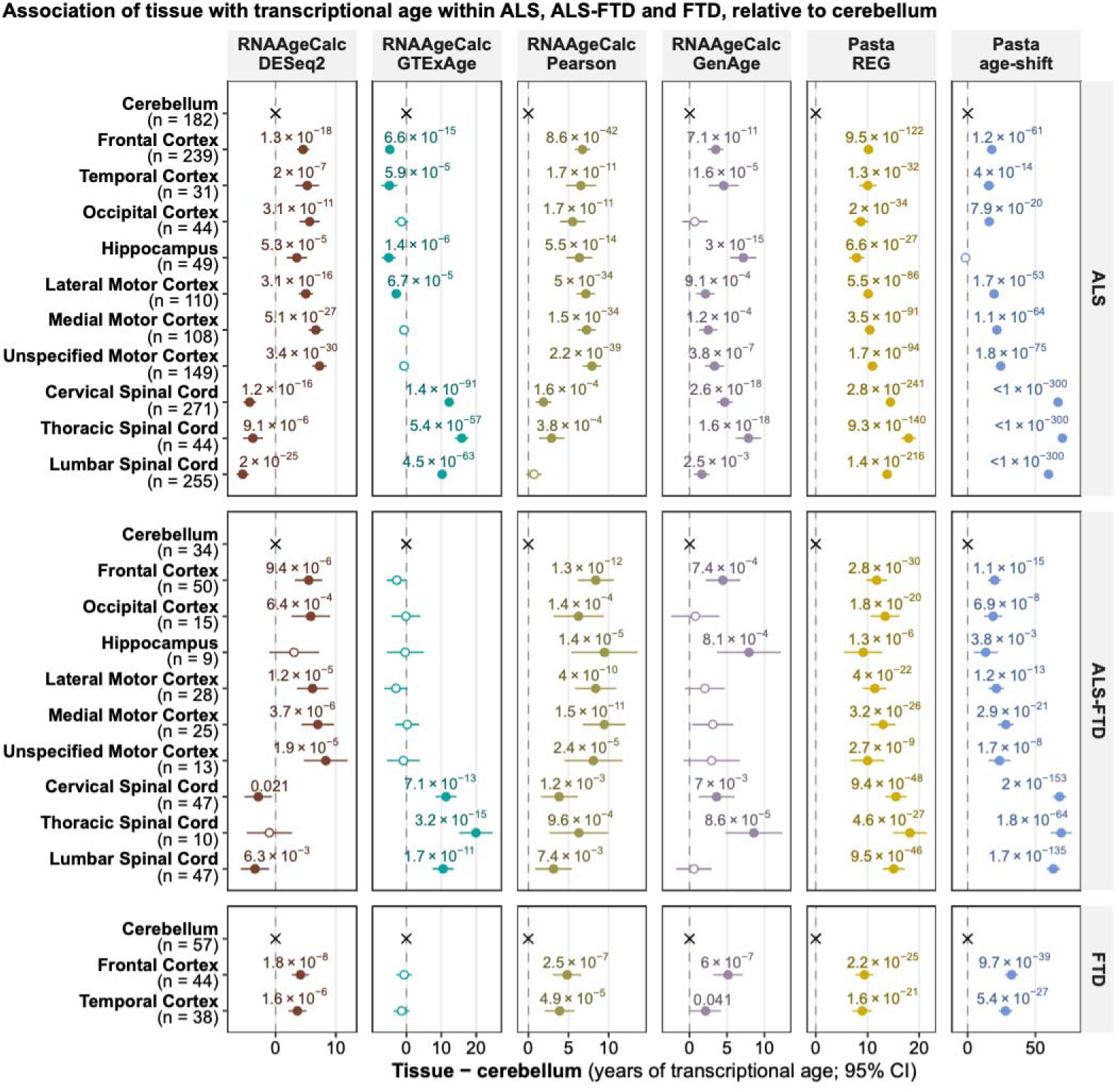
Transcriptional age differences across tissues within ALS, ALS-FTD and FTD. Sections are disease groups (ALS, top; ALS-FTD, middle; FTD, bottom) and rows are tissue regions. Each column is a clock with units of years of transcriptional age or age-score years (Pasta age-shift). Tissue−cerebellum differences were quantified using separate linear mixed models within each group, adjusted for chronological age, library preparation and sex, with a donor random intercept (ALS, 1,482 samples, 378 donors; ALS-FTD, 278 samples, 55 donors; FTD, 139 samples, 60 donors; Methods 2.4). Filled points mark FDR < 0.05 (FDR labeled). FDR correction was performed across tissue contrasts within disease group and clock. Crosses mark cerebellum as the zero reference. n, tissue donors; no more than one sample per donor/tissue. Normal approximation 95% CIs, estimate ± 1.96 × model SE, from restricted maximum likelihood.

Across ALS and ALS-FTD, most tissue contrasts were positive relative to cerebellum for five of the six clocks. Both Pasta models showed higher adjusted transcriptional age across the tested tissues, and predominantly positive contrasts were also observed for the RNAAgeCalc Pearson and GenAge clocks, although fewer GenAge contrasts in ALS-FTD remained significant after FDR correction. RNAAgeCalc DESeq2 showed a distinct regional pattern, with lower estimates in spinal cord and higher estimates in cortical regions relative to cerebellum, consistent with the spinal DESeq2 reductions observed in the disease–control analyses. GTExAge showed a different pattern, with spinal cord estimates significantly higher than cerebellum, whereas most cortical estimates were lower or did not differ significantly. Overall, ALS and ALS-FTD showed similar directional regional patterns, although fewer contrasts reached FDR significance in ALS-FTD.

In FTD, frontal and temporal cortex showed higher adjusted transcriptional age than cerebellum for five of the six clocks, including both Pasta models, with most contrasts remaining significant after FDR correction. GTExAge was again the exception, with neither cortical contrast remaining significant after FDR correction.

### 3.4 *CHIT1* term accounts for the RNAAgeCalc DESeq2 decrease in ALS spinal cord

RNAAgeCalc DESeq2 was the only clock to show significantly lower transcriptional age in ALS cervical and lumbar spinal cord than in controls (Figure 1). We therefore examined the contribution of individual genes to this decrease. *CHIT1* combined a large negative clock coefficient with the largest ALS-associated expression increase among the clock’s genes (Figure 3A). Of all 472 clock genes, *CHIT1* was the only gene to contribute an absolute value greater than 5 transcriptional years to the ALS-control difference (−8.37 years in cervical spinal cord; - 5.34 years in lumbar spinal cord). Because the clock weight is negative, higher *CHIT1* expression in ALS produced a negative contribution to RNAAgeCalc DESeq2 transcriptional age. *CHIT1* expression was significantly higher in ALS than in controls after covariate adjustment in both cervical and lumbar spinal cord, resulting in a significant negative *CHIT1* term difference between ALS and controls. Its negative contribution exceeded the overall decrease at both spinal cord levels and was therefore partly offset by contributions from other genes in the full DESeq2 model. Removing the *CHIT1* term without retraining the model yielded a significantly higher transcriptional age in ALS relative to controls in cervical spinal cord, while the difference in the lumbar spinal cord was no longer significant (Figure 3B).

**Figure 3.**
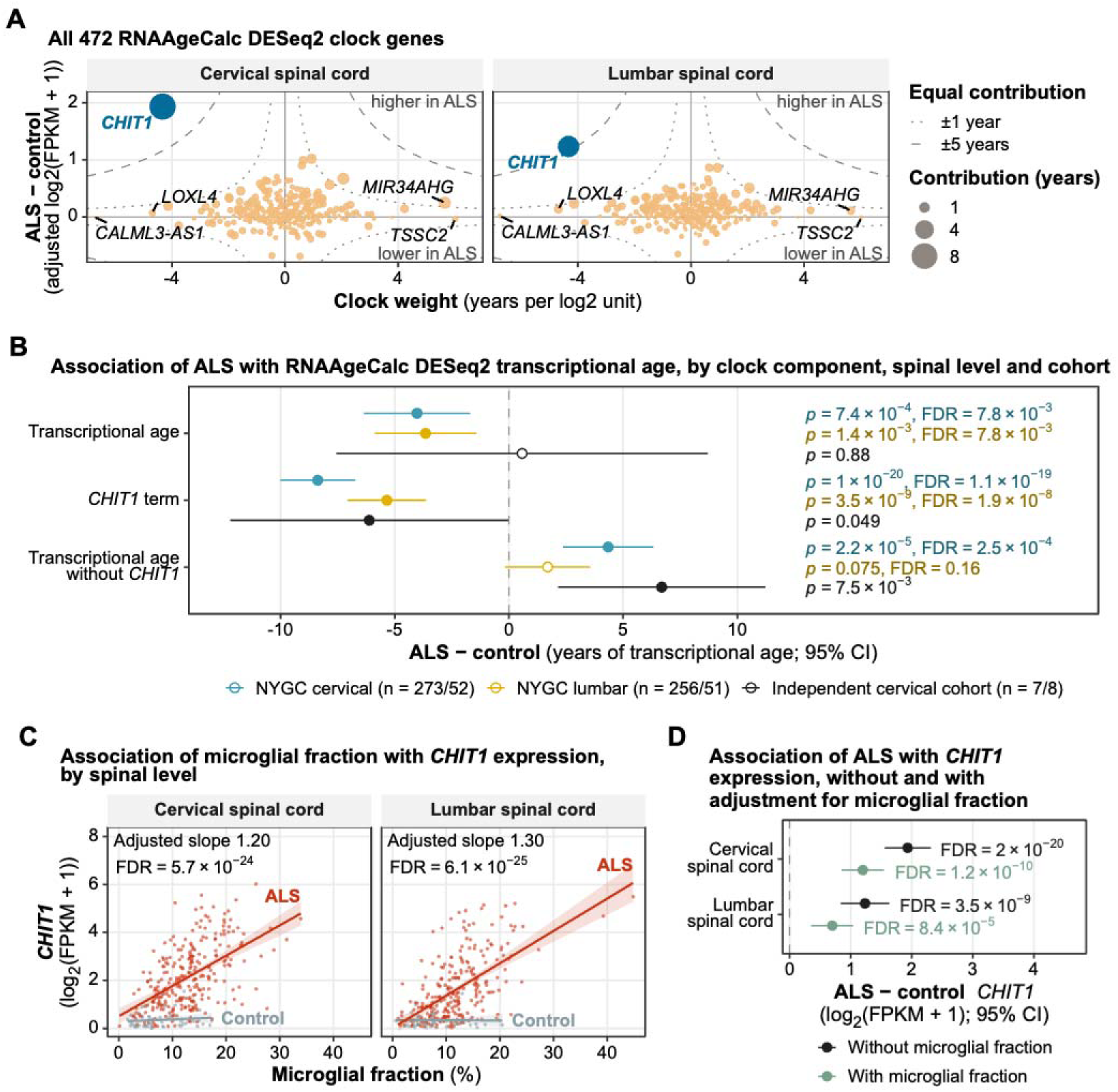
Contribution of *CHIT1* term to RNAAgeCalc DESeq2 transcriptional age models and association with ALS. (A) Clock feature weights versus adjusted ALS−control expression differences across 472 DESeq2 model genes. Point area is the absolute contribution. Curves indicate ±1/±5-year contributions. The four largest absolute weights and *CHIT1* are labeled. (B) ALS differences for the full DESeq2 clock, *CHIT1* term, and remaining gene component (471 genes) in NYGC cervical (blue) and lumbar (yellow) cord and in the independent cervical spinal cord cohort (black). The independent-cohort model was adjusted for age. Filled points mark FDR < 0.05 across tissues within outcome (Methods 2.5), or p < 0.05 for the independent cohort (one test per outcome); n, ALS/controls. (C) *CHIT1* log_2_(FPKM + 1) versus microglial fraction. Lines are unadjusted fits by disease group. Inset annotation indicates slope and FDR for the model of adjusted expression change per 10-percentage-point increase in cell fraction. (D) ALS−control *CHIT1* differences before/after adjustment for microglial fraction. Primary models used age and cord-specific technical covariates (Methods 2.3); models for (C–D) additionally included ALS status. 95% CIs (B, D); FDR across cord levels (C–D).

We next evaluated these findings in an independent cervical spinal cord bulk RNA-seq cohort. The overall ALS-associated decrease in RNAAgeCalc DESeq2 transcriptional age was not replicated. However, the *CHIT1* term was significantly negatively associated with ALS status, whereas DESeq2 transcriptional age without the *CHIT1* term was positively associated with ALS status. Thus, although the overall clock result was not replicated, the independent cohort supported the opposing contribution of *CHIT1* to the DESeq2 transcriptional age estimate.

In ALS spinal cord, *CHIT1* has previously been found to be increased in microglia and macrophages (Steinacker et al. 2018), so we also examined the association between the microglial fraction and *CHIT1* expression. The NYGC ALS Consortium provides estimated cell-type fractions derived from deconvolution of bulk RNA-seq data for a subset of the CNS tissues, including cervical and lumbar spinal cord. We therefore modeled *CHIT1* expression as a function of ALS status with and without adjustment for estimated microglial fraction. Higher microglial fraction was significantly associated with higher *CHIT1* expression in both cervical and lumbar spinal cord (Figure 3C). Adjustment for microglial fraction attenuated the positive association between ALS status and *CHIT1* expression, although an ALS-associated difference remained (Figure 3D). This result indicates that the estimated microglial fraction may partly account for, but not fully explain, the ALS–control difference in *CHIT1* expression.

### 3.5 ALS-associated transcriptional age differences show tissue-specific sensitivity to cell composition

We next investigated whether ALS-associated changes in cell composition contributed to the transcriptional age differences observed across multiple clocks. To address this, we first characterized differences in estimated cell type fractions between ALS and control tissues. After FDR correction, most significant associations were in the cervical and lumbar spinal cord (Supplementary Figure S2). In ALS spinal cord, oligodendrocyte fractions were significantly lower and astrocyte, microglial and lymphocyte fractions were higher; lumbar Schwann-cell and pericyte fractions were also higher.

Six cortical and cerebellar cell fraction comparisons remained significant after FDR correction. In contrast to the spinal cord, ALS frontal cortex showed lower astrocyte and higher oligodendrocyte fractions, and astrocyte fractions were also lower in medial motor cortex. No significant cell fraction differences were observed in unspecified motor cortex.

For cell composition to contribute to an ALS-associated clock difference, the relevant cell fractions must differ between ALS and controls and be associated with the clock estimate. We therefore examined the association between transcriptional age and ALS status before and after adjustment for individual cell type fractions and for overall cell composition in the three tissues showing the strongest ALS-associated differences: cervical spinal cord, lumbar spinal cord, and unspecified motor cortex.

In the spinal cord, adjustment for cell composition improved model fit for all clocks and attenuated several ALS-control differences (Supplementary Figure S3; Figure 4). GTExAge, Pearson, Pasta REG, Pasta age-shift, and cervical GenAge no longer passed FDR correction after adjustment, leaving lumbar GenAge as the only significant positive association. The ALS-associated RNAAgeCalc DESeq2 decreases remained significant.

**Figure 4.**
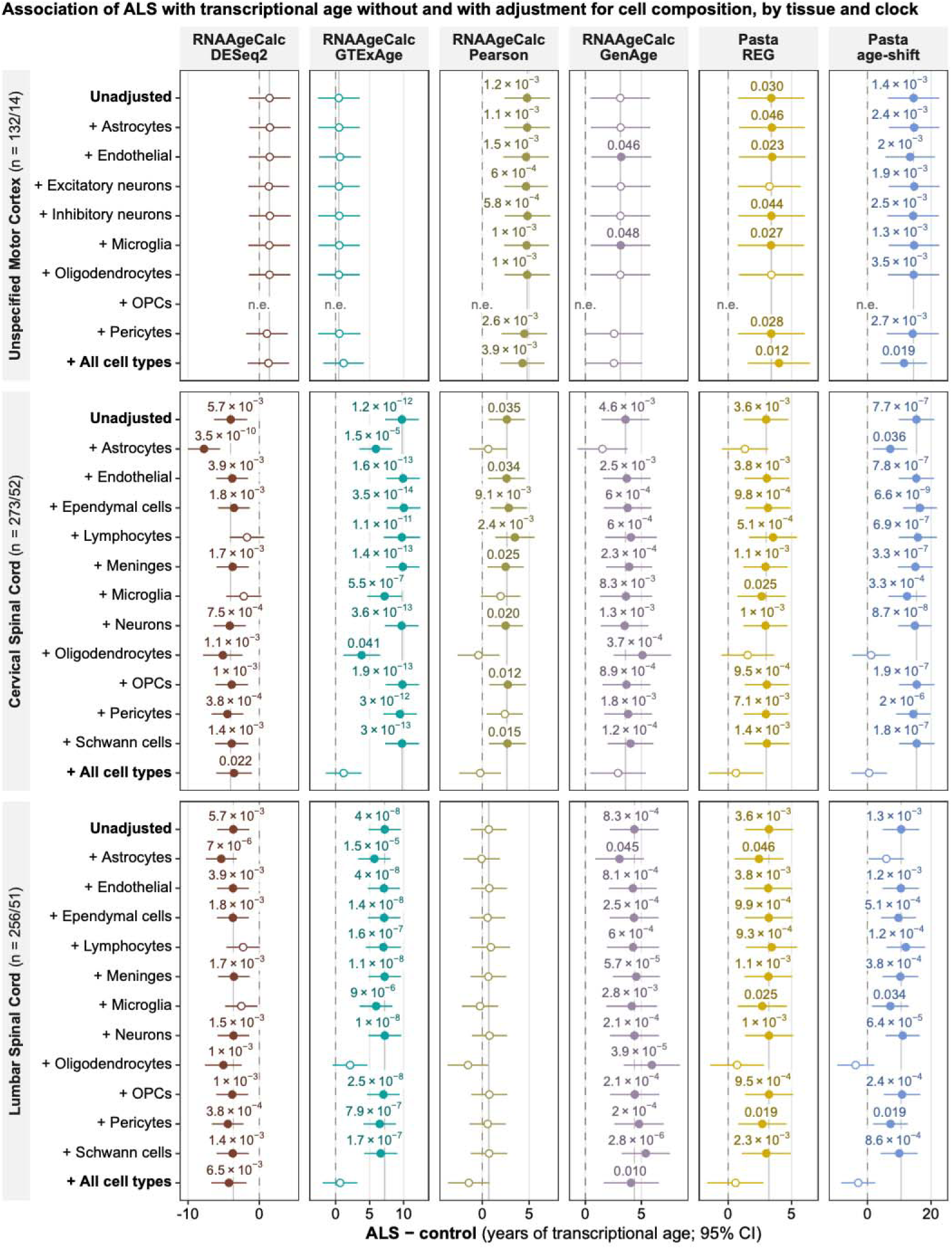
Effect of cell composition on transcriptional age differences. Sections are tissues with at least one FDR-significant ALS-control difference before cell-composition adjustment (n, ALS/control samples). Rows indicate individual cell-type fractions added to the primary model in samples with composition data; “Unadjusted” is the primary model, and “All cell types” includes all fractions except the reference cell type; n.e., fraction was 0 in every sample (not estimable). Each column shows the ALS–control estimate and 95% CIs for a clock with units of years of transcriptional age or age-score years (Pasta age-shift). Filled points mark FDR < 0.05 across the eight tissues with cell composition estimates within clock and adjustment. Vertical grey line marks the unadjusted estimate.

When cell fractions were considered individually, oligodendrocyte adjustment attenuated the spinal ALS-control differences for GTExAge, Pearson, Pasta REG, and Pasta age-shift, but strengthened the GenAge differences in both cord regions. After adjustment for oligodendrocyte fraction, three of six significant cervical and three of five significant lumbar associations no longer passed FDR correction. Astrocyte adjustment similarly attenuated most clock differences, with three cervical and one lumbar association losing FDR significance, but the RNAAgeCalc DESeq2 decreases became stronger after adjustment. Adjustment for lymphocyte or microglial fraction had comparatively little effect on most clocks; however, microglial adjustment attenuated the cervical Pearson increase, and adjustment for either microglia or lymphocytes attenuated the RNAAgeCalc DESeq2 decreases in both spinal cord regions, consistent with the relationship between microglial abundance and *CHIT1* expression (Results 3.4). Overall, spinal cord cell composition contributed substantially to the ALS-associated differences in transcriptional age. Among the cell types examined, oligodendrocyte and astrocyte fractions produced the greatest attenuation of ALS-associated transcriptional age differences across most clocks, whereas lymphocyte and microglial fractions had a greater relative effect on RNAAgeCalc DESeq2.

In unspecified motor cortex, inclusion of cell composition also improved the model fit for all clocks, though the partial F-test for the DESeq2 clock was only nominally significant (p = 0.05). In contrast to the spinal cord, the significant positive associations generally retained their direction and significance after adjustment for overall cell composition. Individual cell type adjustment showed no consistent pattern of attenuation across clocks. In GenAge, adjustment for endothelial or microglial fraction strengthened the positive ALS–control difference, which then passed FDR correction. Conversely, adjustment for excitatory neuron or oligodendrocyte fraction attenuated the Pasta REG difference, which no longer passed FDR correction. Overall, both joint and individual cell type adjustments had less consistent effects on ALS-associated transcriptional age differences in unspecified motor cortex than in the spinal cord.

### 3.6 RNAAgeCalc DESeq2 and GTExAge transcriptional age acceleration are associated with ALS disease duration

Having examined the contribution of cell composition to transcriptional age estimates, we next investigated whether AA was associated with disease duration in ALS and ALS-FTD. Among the five clocks that generally estimated higher transcriptional age in ALS, only GTExAge showed significant associations between AA and disease duration in ALS. Higher GTExAge AA in the occipital cortex and cervical and thoracic spinal cord was associated with an increased hazard of death, indicating shorter ALS duration (Figure 5A). No associations in ALS-FTD remained significant after FDR correction.

**Figure 5.**
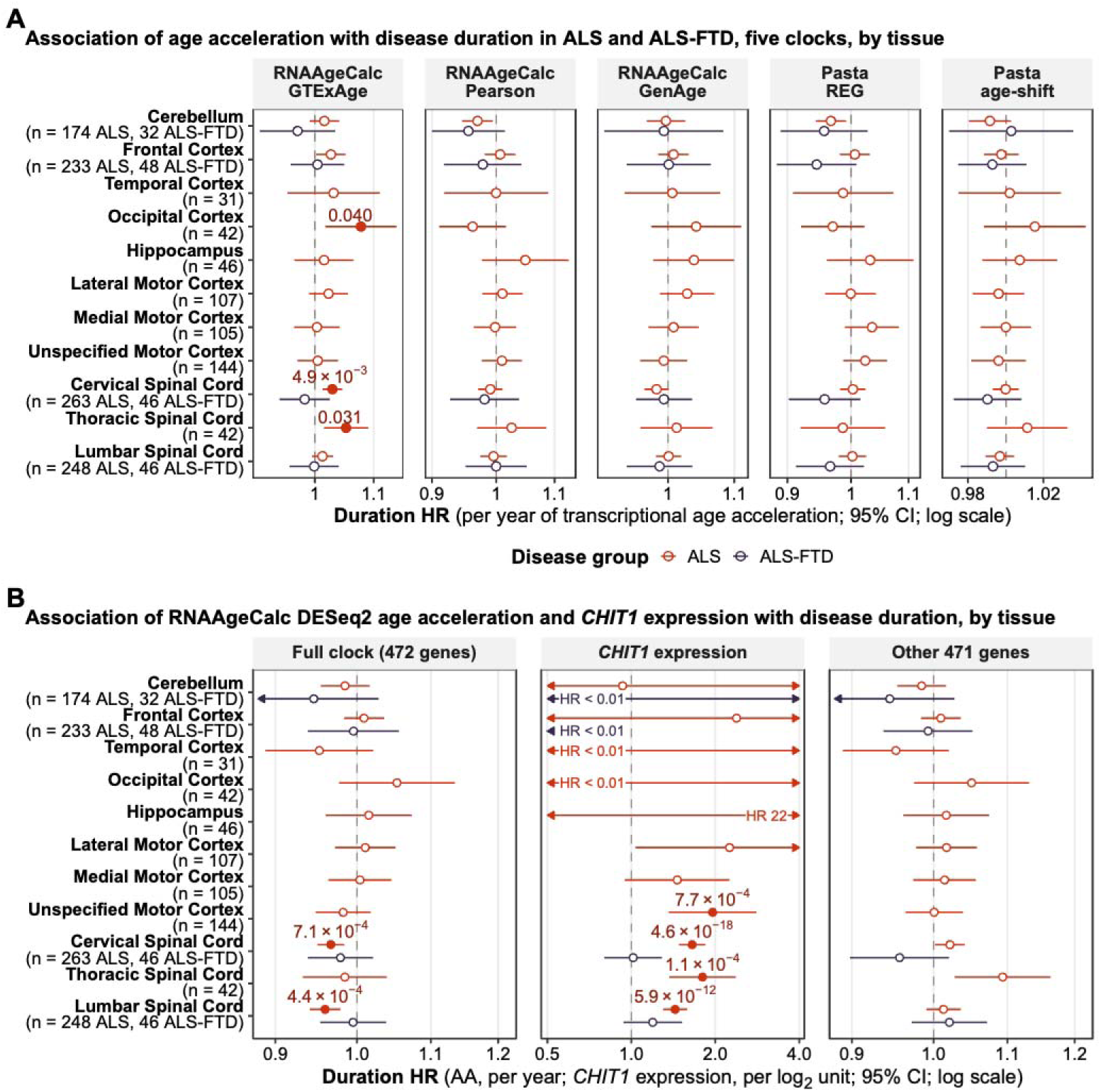
Association between age acceleration (AA) and disease duration. (A) Hazard ratios (HR) and 95% CIs per year of transcriptional AA for GTExAge, Pearson, GenAge and Pasta clocks. (B) Hazard ratios and 95% CIs for the RNAAgeCalc DESeq2 full-clock (472 genes; first panel) and remaining gene component (471 genes; third panel) per AA year. *CHIT1* expression HRs per log_2_(FPKM + 1) unit (second panel). AA is the residual of transcriptional age adjusted for chronological age and technical covariates. Arrows mark truncated CIs. Off-scale HRs are labelled. Cox models were adjusted for library preparation and sex. HR < 1 indicates a lower hazard of death and longer disease duration; HR > 1 indicates a higher hazard of death and shorter disease duration. Filled labelled points mark FDR < 0.05 across tissues within group/clock/exposure. Log scales; n, donors.

In contrast, higher DESeq2 AA was associated with longer ALS duration in cervical and lumbar spinal cord (Figure 5B). Higher *CHIT1* expression was associated with shorter ALS duration at all cord levels and in unspecified motor cortex. After removing *CHIT1* from the DESeq2 model, the association between higher AA and shorter ALS duration in the thoracic spinal cord did not meet the FDR significance threshold (FDR = 0.053). No duration associations in ALS-FTD remained significant after FDR correction.

### 3.7 No clock differs by *C9orf72* repeat expansion status

We also examined whether transcriptional age differed by *C9orf72* repeat expansion status. Across all tested tissues and clocks, no differences between *C9orf72* expansion carriers and non-carriers remained significant after FDR correction. This remained the case after adjustment for estimated cell-type composition in tissues with available deconvolution data (Supplementary Figure S4).

## 4. Discussion

In this study, we characterized tissue-specific differences in transcriptional age across ALS, ALS-FTD, and FTD using six transcriptional aging clocks. Disease-associated differences varied across tissues and clocks, with the strongest ALS and ALS-FTD differences occurring in the spinal cord. Significant changes were also identified in the unspecified motor cortex in ALS and in the frontal and temporal cortices in FTD. We further examined the contribution of disease-associated gene expression and cell composition to these differences and tested whether transcriptional AA was related to disease duration and whether transcriptional age differed by donor *C9orf72* repeat expansion status. Together, these analyses show that transcriptional aging estimates in ALS and FTD are strongly dependent on the clock and tissue examined and may reflect disease-associated gene expression and cellular composition in addition to aging-related processes.

Although most clocks correlated significantly with chronological age across tissues, their performance was not uniform. Correlations were higher in controls than in patients for 54 of 66 clock–tissue combinations, despite a greater number of significant correlations in the larger pooled patient group, emphasizing that statistical significance alone does not indicate stronger age prediction. Differences in calibration and predicted age range, including compression of RNAAgeCalc DESeq2 estimates, further support interpreting disease-associated differences in the context of individual clock performance.

### 4.1 Gene-level contributions to the RNAAgeCalc DESeq2 decrease

The direction of ALS–control differences in the spinal cord depended on the clock. Most of the clocks estimated significantly higher transcriptional age in ALS, whereas RNAAgeCalc DESeq2 estimated lower age. Gene-level decomposition and cell composition adjustment traced these opposite shifts to two related sources. A single gene, *CHIT1*, was the primary contributor to the DESeq2 decrease in ALS. In our analysis, estimated microglial fraction provided a link between the gene-level and cell composition effects, contributing to the increase in *CHIT1* expression. In addition, adjustment for either microglial or lymphocyte fractions attenuated the ALS-associated decrease in DESeq2 transcriptional age.

*CHIT1* encodes chitotriosidase, which is expressed by cells of the monocyte/macrophage lineage and associated with innate immunity and inflammation (Steinacker et al. 2018). In postmortem ALS spinal cord, chitotriosidase was found mainly in the lateral and anterior corticospinal tracts, where it colocalised with subpopulations of IBA1-positive microglia and CD68-positive macrophages (Steinacker et al. 2018). In the brain, chitotriosidase staining was sparse and much weaker (Steinacker et al. 2018). Chitotriosidase levels were also raised in ALS cerebrospinal fluid (Steinacker et al. 2018) and correlated with the rate of disease progression (Thompson et al. 2019; Steinacker et al. 2018).

Because of its large negative weight in the DESeq2 clock, the marked increase in *CHIT1* expression in ALS spinal cord produced a negative contribution larger than the net clock decrease, with other genes partially offsetting the effect of *CHIT1* in the full clock. Higher estimated microglial abundance was associated with, and partially contributed to, the *CHIT1* expression difference. In the independent cervical dataset, *CHIT1* was again the largest negative contributor, but other gene contributions offset it and the full DESeq2 model did not show a significant ALS-associated difference in transcriptional age. These findings show that a single disease-associated gene can strongly influence a multigene clock, underscoring the need for gene-level decomposition before interpreting transcriptional age differences as biological aging.

### 4.2 Cell-associated contributions to transcriptional aging differences

Cell composition also emerged as a major contributor to the clocks that demonstrated higher transcriptional age in ALS spinal cord. Inclusion of cell composition as a covariate attenuated ALS spinal cord differences in all clocks except the lumbar GenAge, which remained significant. Cell composition is a plausible contributor to an ALS-control difference in transcriptional age when the estimated cell type fraction differs by disease status and is associated with the clock. In the NYGC data, ALS spinal cord had lower estimated oligodendrocyte fractions and higher astrocyte, microglial, and lymphocyte fractions than non-neurological control spinal cord. These differences were consistent with the glial activation and reduced oligodendrocyte-associated expression reported in an earlier NYGC spinal-cord cohort (Humphrey et al. 2023). Consistent with a compositional contribution, adjustment for oligodendrocyte fraction alone attenuated most of the ALS-control clock differences, whereas astrocyte adjustment preferentially attenuated the GenAge and cervical Pasta REG increases.

Because bulk tissue deconvolution relies on reference expression signatures, inferred cell-type fractions can be influenced by both true compositional differences and alterations in cell type-specific expression. Deconvolution alone cannot reliably distinguish these sources (Wolfram-Schauerte et al. 2025). In the ALS consortium resource, the cell composition estimates were derived from bulk expression using tissue-matched single-cell references, with the single-nucleus reference samples restricted to non-neurological donors. Because reference-based deconvolution assumes that reference expression signatures adequately represent the target tissue, ALS-associated reactive states absent from the reference could influence the inferred fractions. Thus, the estimated differences may reflect true changes in cell abundance, altered cell state, or both. This limitation was also recognized in the NYGC study, which noted that bulk RNA-seq cannot distinguish true composition changes from cell-state shifts (Humphrey et al. 2026).

In the case of glial fractions, both interpretations are biologically plausible. Mature oligodendrocyte degeneration has been demonstrated in SOD1-mutant ALS mice (Kang et al. 2013), whereas human postmortem studies have identified reactive oligodendrocyte, microglial, and astrocyte states (Masrori et al. 2025; Zelic et al. 2025; Georgopoulou et al. 2025). Consequently, attenuation of ALS-associated clock differences after adjustment for deconvolved cell composition supports the sensitivity of these clocks to cell-associated signals but does not establish whether those signals reflect changes in cell abundance, cellular state, or both.

Importantly, sensitivity to cell composition does not necessarily make these transcriptional aging signals biologically uninformative or distinguish disease-related remodeling from biological aging. In a recent whole blood study of epigenetic aging in ALS, apparent ALS–control differences were eliminated after adjustment for white blood cell proportions, showing that the unadjusted signals were strongly dependent on immune cell composition (Merbaum et al. 2025). As discussed by Merbaum *et al*., such composition-dependent signals are not necessarily biologically uninformative, as changes in immune cell populations may themselves reflect aging processes (Merbaum et al. 2025). This distinction is also relevant to transcriptional aging, as immune cell shifts are features of immunosenescence (Castro et al. 2024; von Leden et al. 2017) and age-related CNS changes include alterations in glial abundance and activation state (Labarta-Bajo and Allen 2025; Tylek and Basta-Kaim 2025). Accordingly, attenuation of spinal transcriptional age differences after composition adjustment indicates sensitivity to disease-associated cell-type signals while leaving open how those signals relate to ALS pathobiology, biological aging, or processes shared by both.

Beyond these ALS-specific findings, the regional distribution of transcriptional age differences broadly aligned with the clinical and anatomical features of the ALS-FTD spectrum. In ALS and ALS-FTD, the strongest disease-associated differences occurred in the spinal cord, whereas in FTD, significant differences were observed in frontal and temporal cortices. Within-disease comparisons similarly showed higher transcriptional age in frontal and temporal cortex relative to cerebellum in FTD across most clocks. Although the smaller number of evaluable tissues and samples in ALS-FTD and FTD limits direct comparisons between disease groups, these findings suggest that transcriptional aging signatures vary across anatomically relevant regions of the ALS-FTD spectrum rather than representing a uniform CNS-wide shift.

### 4.3 Disease duration and *C9orf72*

Associations with clinical features further illustrate that transcriptional clocks can capture distinct aspects of ALS biology. Higher DESeq2 AA was associated with longer disease duration, but this association was strongly influenced by *CHIT1*, with higher spinal *CHIT1* expression corresponding to shorter duration. This direction is consistent with cerebrospinal fluid chitotriosidase levels that correlate with faster progression (Thompson et al. 2019) and were associated with survival in one cohort (Thompson et al. 2019). By contrast, higher GTExAge AA was associated with shorter disease duration in occipital cortex, cervical and thoracic spinal cord, which is directionally consistent with previous methylation studies linking greater AA to earlier onset and shorter disease duration in *C9orf72* expansion carriers (Zhang et al. 2017) and earlier onset and shorter survival in ALS more broadly (Zhao et al. 2024; Zhang et al. 2020). However, this relationship has not been consistent across molecular aging studies, as a subsequent large methylation-based analysis found no association between epigenetic AA and ALS survival (Merbaum et al. 2025).

We found no evidence that tissue transcriptional age differed by *C9orf72* repeat expansion status after FDR adjustment. This contrasts with the recent whole blood methylation study by Merbaum and colleagues, in which *C9orf72* expansion carriers showed higher DunedinPACE and shorter DNAm-estimated telomere length after adjustment for white blood cell composition (Merbaum et al. 2025). The discrepancy may reflect differences in molecular data type, tissue, clock construction, and statistical power rather than conflicting evidence about aging in *C9orf72* ALS. Indeed, the methylation study itself found no *C9orf72*-associated difference for the Horvath, PCGrimAge, or pan-mammalian clocks, demonstrating that even within a single molecular layer the apparent aging phenotype depends on the clock used. Together with the divergent tissue and clinical associations observed across transcriptomic clocks in the present study, these findings argue against interpreting transcriptional AA as a single, clock-independent measure of biological aging.

### 4.4 Limitations

Several limitations constrain the interpretation and generalizability of these findings. MuSiC cell composition estimates were unavailable for ALS-FTD and FTD and for several regions, including thoracic spinal cord, occipital cortex, and hippocampus, preventing composition-adjusted analyses in these groups and tissues. Moreover, cell fractions were estimated from bulk RNA-seq and may reflect both differences in cellular abundance and disease-associated changes in cell state, as discussed above. Independent validation was also limited by the small cervical spinal cord cohort, which included only seven ALS and eight control donors. The cross-sectional, postmortem design also limits our ability to distinguish aging-related changes from changes occurring as a consequence of disease, and sample availability differed substantially across tissues and disease groups, particularly for ALS-FTD and FTD. Finally, the RNAAgeCalc brain models were trained on all GTEx CNS tissues, which may limit their calibration and biological specificity. More broadly, differences in performance and direction across clocks indicate that transcriptional age estimates are dependent on clock construction and should not be interpreted as direct measures of biological aging.

## 5. Conclusions

In this study, we characterized tissue-specific differences in transcriptional age across the ALS– FTD spectrum using six transcriptomic aging clocks and investigated how these signals relate to disease-associated gene expression, cellular composition, clinical duration, and *C9orf72* repeat expansion status. Disease-associated differences in transcriptional age localized to clinically relevant tissues but varied in direction and magnitude across clocks. In ALS, the strongest differences were localized to the spinal cord, where RNAAgeCalc DESeq2 estimated lower transcriptional age, whereas the other clocks estimated higher age when significant. Gene-level decomposition identified *CHIT1* as the primary contributor to the DESeq2 decrease, emphasizing the importance of gene-level model decomposition when interpreting transcriptional clock estimates. Adjustment for estimated spinal cord cell composition substantially attenuated several of the ALS-associated increases observed with other clocks. Associations with disease duration were likewise clock-dependent, and no differences by *C9orf72* repeat expansion status remained significant after FDR correction. Together, these findings suggest that transcriptional aging signatures across the ALS–FTD spectrum reflect interactions among disease-associated gene expression, regional vulnerability, cellular remodeling, and the molecular features captured by individual aging clocks, and should not be interpreted as direct, clock-independent measures of biological aging.

## Data Availability

All data produced in the present study are available upon reasonable request to the authors

https://www.ncbi.nlm.nih.gov/geo/query/acc.cgi?acc=GSE137810

https://trace.ncbi.nlm.nih.gov/Traces/?view=study&acc=SRP064478

https://zenodo.org/records/18989237

https://zenodo.org/records/20090702

## Declarations

### Ethics approval and consent to participate

This study used publicly available, deidentified data and did not enroll participants. Ethics approval and informed consent for the source studies are described in the respective publications.

### Consent for publication

Not applicable.

### Availability of data and materials

The datasets analysed in this study are publicly available through GEO (GSE137810), Zenodo (18989237 and 20090702), and the Sequence Read Archive (SRP064478), as detailed in Supplementary Table S1. Custom scripts generated for this study will be made available (in a public repository upon publication/upon reasonable request).

### Authors’ contributions

Author Contributions: Conceptualization, T.Z. and M.M.H.; methodology, T.Z.; formal analysis, T.Z.; writing—original draft preparation, T.Z.; writing—review and editing, T.Z., M.M.H., G.W. and V.V.B. All authors contributed to the interpretation of the results. All authors have read and agreed to the published version of the manuscript.

## Acknowledgements

The authors gratefully acknowledge the investigators and participants whose contributions made the publicly available datasets used in this study possible, with particular thanks to Dr. Jack Humphrey of the Icahn School of Medicine at Mount Sinai.

## Supplementary material

Supplementary Table S1. and Figures S1–S4 follow.

## Supplementary tables

**Table S1.** Data sources, algorithms and software.

| Category | Resource | Accession or version |
|---|---|---|
| Data source | NYGC bulk RNA-seq | GSE137810; Zenodo 18989237 |
| Data source | NYGC donor/sample metadata and MuSiC estimates | Zenodo 20090702 (ST1, ST3, ST7) |
| Data source | Independent cervical bulk RNA-seq | SRP064478 (recount3) |
| Algorithm | RNAAgeCalc DESeq2 brain | RNAAgeCalc 1.22.0 |
| Algorithm | RNAAgeCalc GTExAge brain | RNAAgeCalc 1.22.0 |
| Algorithm | RNAAgeCalc Pearson brain | RNAAgeCalc 1.22.0 |
| Algorithm | RNAAgeCalc GenAge brain | RNAAgeCalc 1.22.0 |
| Algorithm | Pasta REG | pasta 0.0.0.9000 |
| Algorithm | Pasta age-shift | pasta 0.0.0.9000 |
| Software | Python | 3.11.17 |
| Software | R | 4.5.3 |

All clock predictions used fixed published weights. MuSiC cell-type proportions were supplied in consortium ST7. NYGC resource, (Humphrey et al. 2026); RNAAgeCalc, (Ren and Kuan 2020); Pasta, (Salignon et al. 2026); independent bulk cohort,  (Brohawn et al. 2016; Wilks et al. 2021).

## Supplementary figures

**Figure S1.**
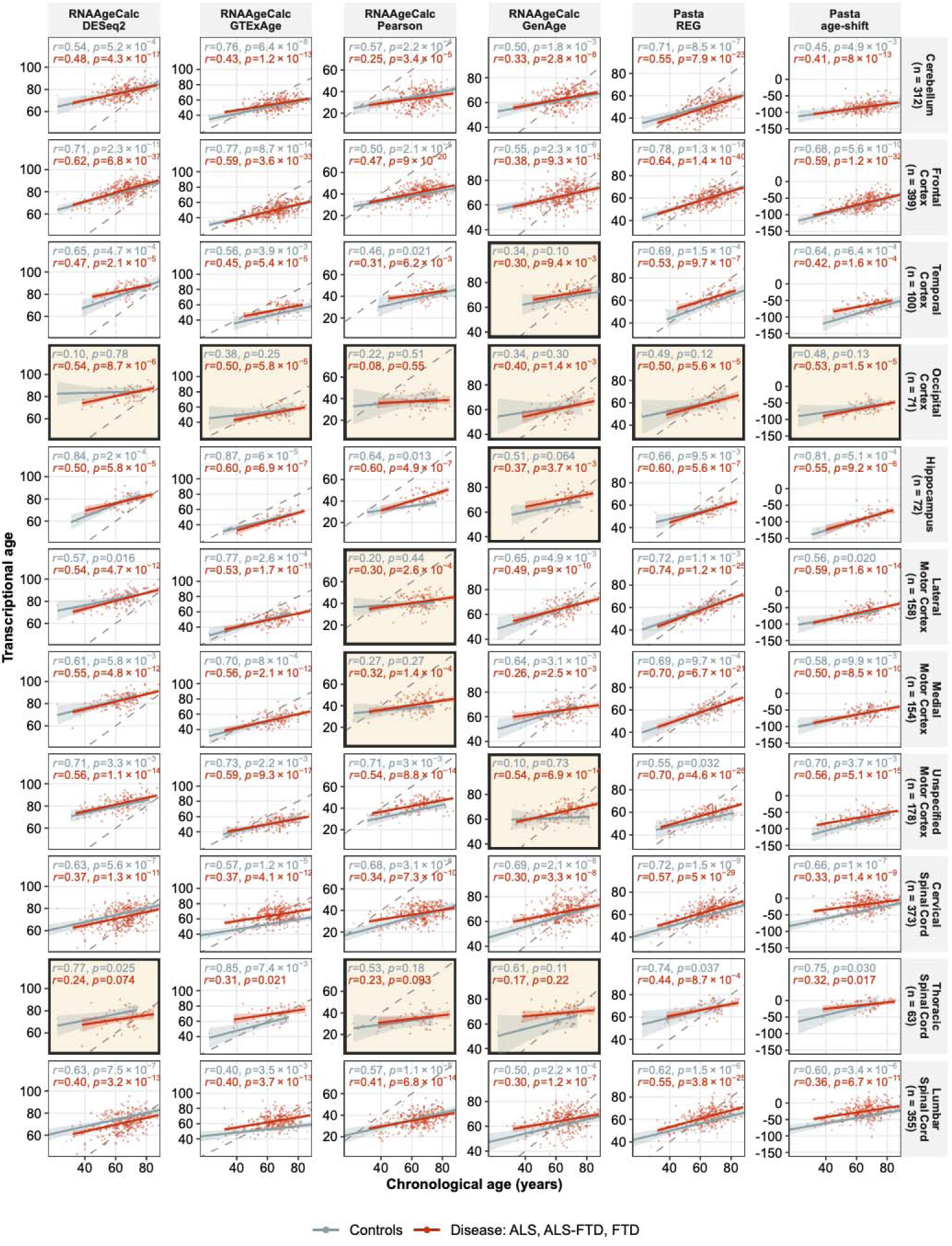
Six clocks versus chronological age. Columns are clocks; rows are tissues. Points and linear fits with 95% CIs are shown separately for controls (grey) and patients (red; ALS, ALS-FTD and FTD pooled). Inset values are Pearson’s r and p within controls (grey, top line) and within patients (red, lower line). Tinted and outlined panels indicate p ≥ 0.05 in at least one group. Dashed identity line except Pasta age-shift. All analysis-set samples.

**Figure S2.**
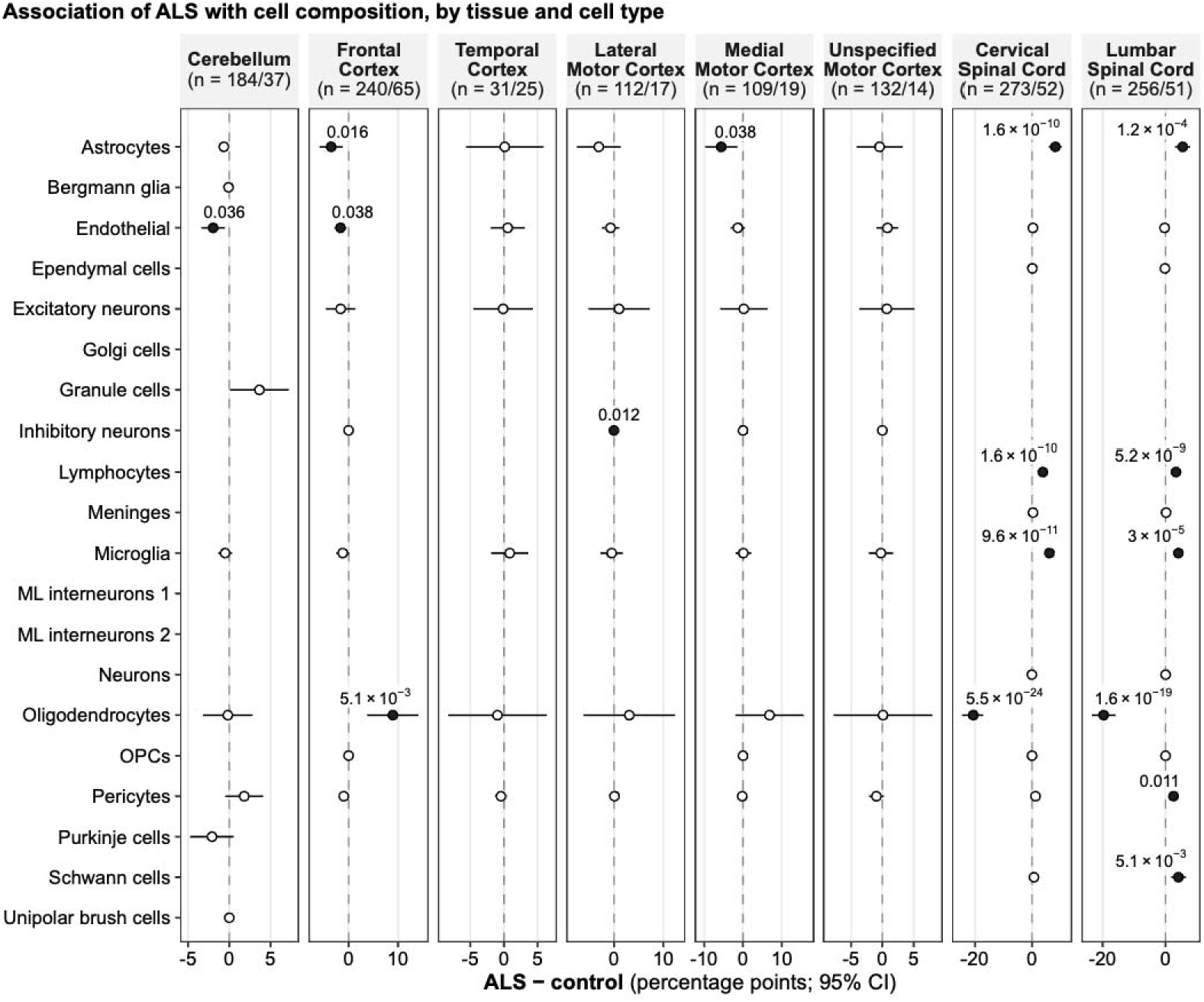
ALS−control differences in cell composition, by tissue and cell type. Rows include every ungrouped MuSiC cell type in the eight tissues’ single-cell references. Columns are tissues with units of percentage-point differences in estimated cell composition (n, ALS/controls). Linear models were adjusted for age and tissue-specific technical covariates (Methods 2.3). Filled points indicate FDR < 0.05 across all 67 tests (FDR labelled). Blank line indicates that the cell type is absent from the tissue’s reference or fraction 0 in every sample (not estimable). ML, molecular layer; OPCs, oligodendrocyte precursor cells.

**Figure S3.**
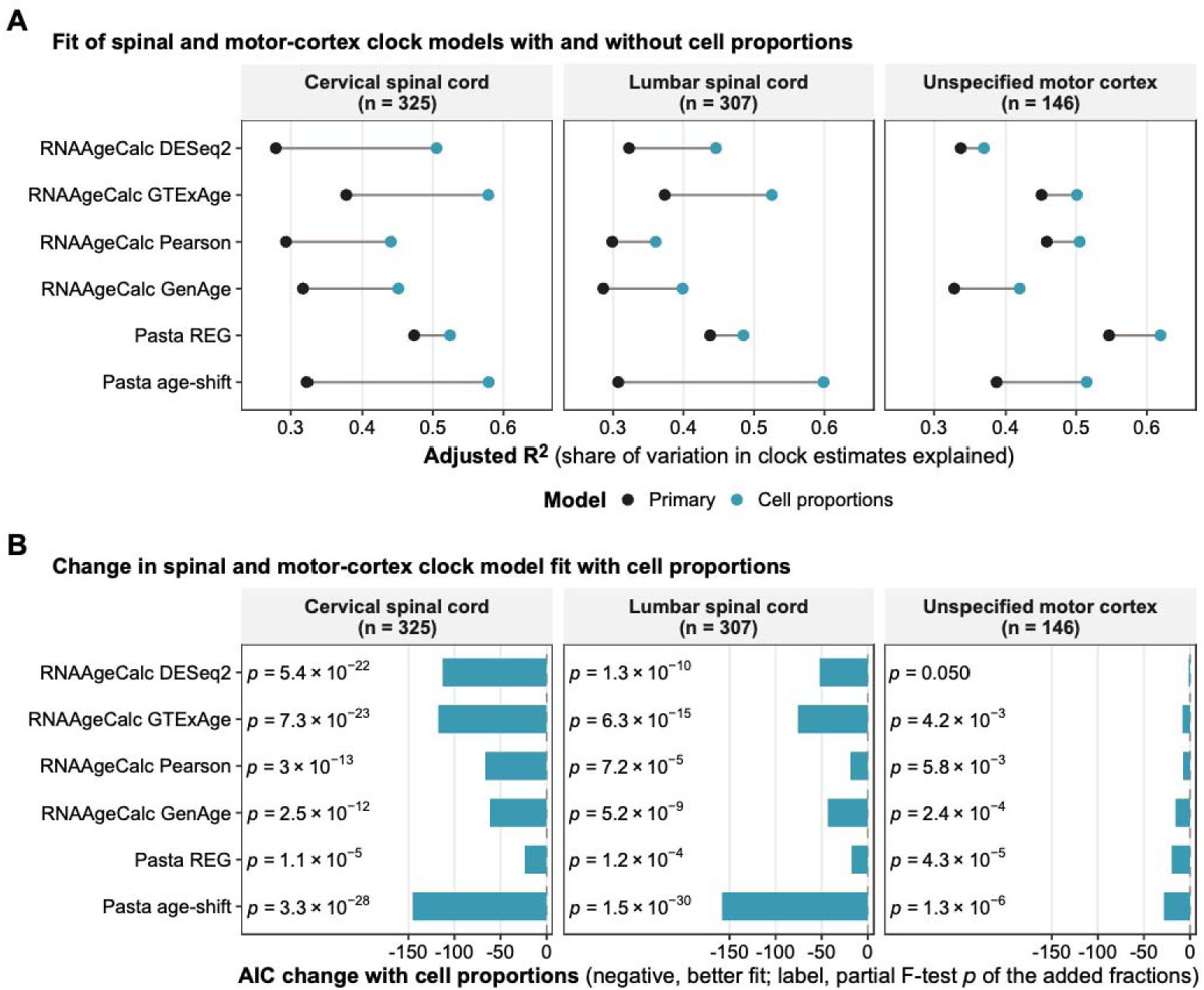
Spinal and unspecified motor-cortex clock models fit with and without cell proportions. Linear models of each clock on ALS status, age and tissue-specific technical covariates (Methods 2.3), fitted on the same ALS and control samples, with and without all variable cell fractions except the reference fraction. (A) Adjusted R^2^. (B) Change in Akaike information criterion (AIC) with cell proportions (negative, better fit). Labels indicate partial F-test p for the added fractions. n, samples.

**Figure S4.**
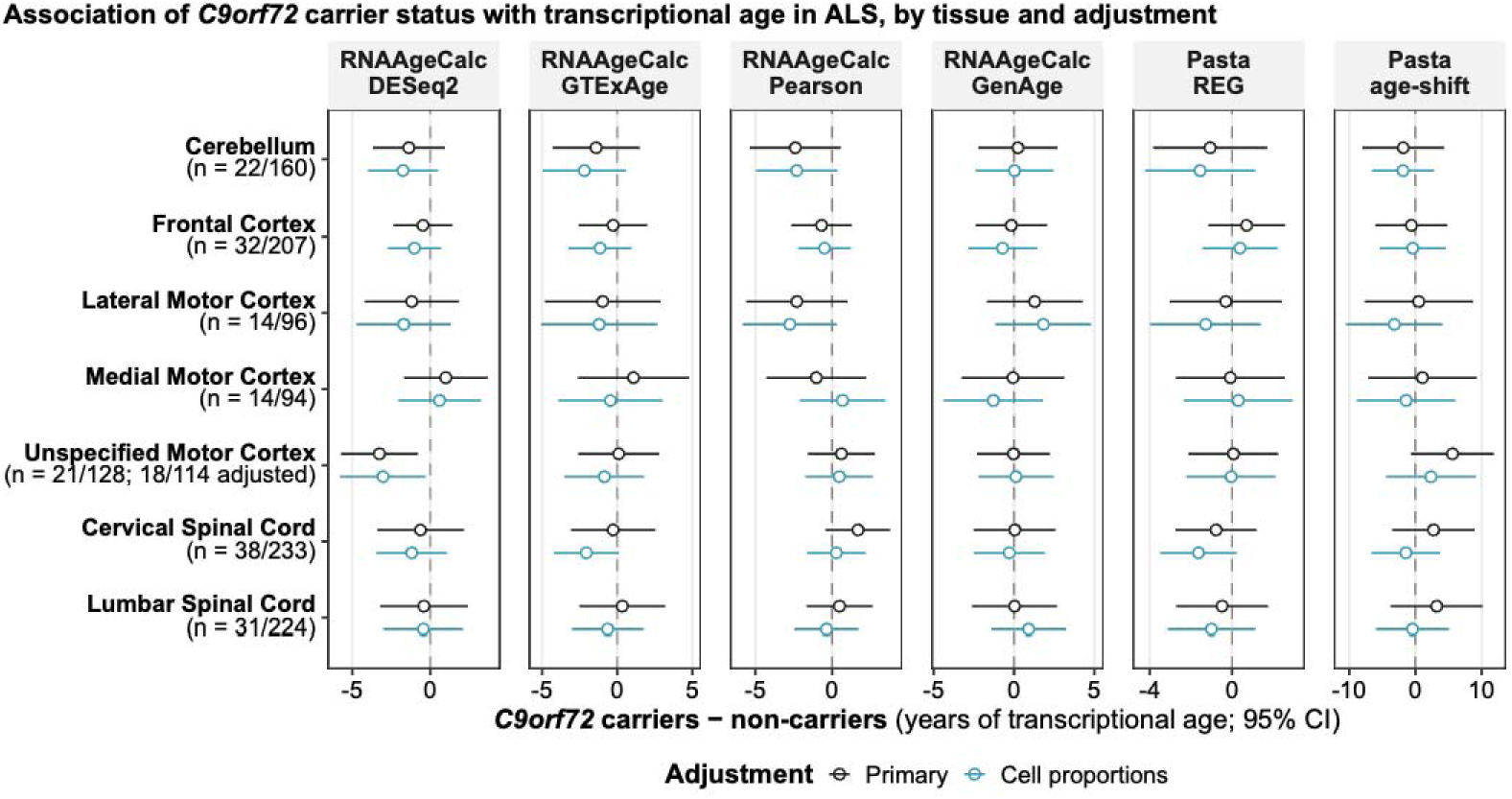
Associations between *C9orf72* repeat expansion status in ALS and transcriptional age across six transcriptomic clocks, by tissue. Carrier−non-carrier differences before (black) and after (blue) cell-composition adjustment. Linear models included library preparation, sex and chronological age, the consortium’s within-ALS covariates (Humphrey et al. 2026) excluding age at onset. Units are years of transcriptional age or age-score years (Pasta age-shift). Filled points indicate an FDR < 0.05 across tissues within clock/adjustment; n, carriers/non-carriers (adjusted n where different).

